# Immunological variables and tumor types influence one-year survival probability in cancer patients: A comprehensive analysis using logistic regression and decision tree models

**DOI:** 10.1101/2023.10.25.23297566

**Authors:** Alvaro López Malizia

## Abstract

The present study aimed to explore immunological variables associated with survival, TP53 gene expression, and primary diagnosis in patients with cancer. Based on these variables, logistic regression and decision tree models (lightGBM) were used to model the probability of one-year survival of patients following their initial diagnosis. Logistic regression revealed the significance of primary diagnosis categories such as Malignant Melanoma, Ovarian Cancer, and Glioblastoma as predictor variables. For the classification model, in addition to these tumor types, variables related to the immune system were also found to be important, including tumor cell percentage, stromal cell percentage, lymphocytes, and necrotic cells. In addition, unsupervised classification techniques were employed to explore the numerical dataset. For this methodology, the best clustering cohesion was observed with two groups determined using different algorithms. The clusters generated by k-means and DBSCAN exhibited differences in the proportion of infiltrating lymphocytes, neutrophils, and monocytes.

## 1. Introduction

Cancer is a group of diseases characterized by uncontrolled division of cells in the body. This abnormal cell division is a result of multiple factors, both genetic and environmental, and can originate in any organ or cellular tissue. In many cases, disease progression is marked by the migration or metastasis of abnormal cells to other parts of the organism. Patient mortality is attributed to the ability of these cells to destroy the tissues in which they appear, disrupting the body’s functions or weakening the individual against other pathologies.

Different types of cells coexist within the tumor tissue. Neutrophils, monocytes, and lymphocytes are immune system cells that can be recruited to tissues and recognize cancer cells. As a result, a balance is established between the immune response and tumor growth. Numerous studies have demonstrated that lymphocyte infiltration into the tumor correlates with a better immune response and overall survival^1–3^. In addition to these cells, the proportions of normal tissue, stromal cells, and damaged tissue (percentage of necrotic or dead cells) is considered. Stromal cells are connective tissue cells that support and coat various organs. Unlike the tumor cells, the stromal cells are normal. The transition from a normal cell to a pro-tumor cell involves numerous internal processes. Some genes protect against transformation and are linked to apoptosis, cell division, and cellular senescence. One such gene that encodes a protein is TP53 (p53 protein).

### 1.2. p53 Protein - TP53 Gene

p53 is a protein present in the nucleus that controls cell division and apoptosis. It is encoded by a single gene (TP53) and belongs to a family of tumor suppressor proteins. p53 is of great importance, as it is altered in the majority of tumors ^4,5^.

### 1.3. Lymphocytes

Lymphocytes are a type of cell belonging to the immune system, characterized by a spherical morphology, are generally smooth, and have little cytoplasm relative to the nucleus. Phenotypically, they are characterized by the expression of the surface markers CD45 and CD3 for T lymphocytes and CD19, CD20, or CD21 for B lymphocytes. Functionally, they play a significant role in adaptive immunity, which is characterized by antibody synthesis (B lymphocytes) or cytotoxic responses (CD8+ T lymphocytes).

### 1.4. Neutrophils

Neutrophils play a significant role in innate immune response and often constitute the first cellular response to bacterial and fungal infections. They have a short lifespan and continuously circulate in the bloodstream. They have a rough morphology and are characterized by the generation of a respiratory burst as a mechanism of cell lysis. They presented the membrane markers CD45+ and CD11b+. Their presence in the tumor tissue, associated with the immune response, affects metastasis, prognosis, and the type of treatment to be developed ^6,7^.

### 1.5. Necrotic Tissue

When cells die, they disintegrate in an orderly manner, preventing the release of their cytoplasmic content into the extracellular medium through the process of apoptosis. This organized type of cell death differs from necrosis, where the cell ruptures and its contents are released into the extracellular medium.

### 1.6. Monocytes

Unlike the cell types mentioned earlier, monocytes have a higher degree of cellular pluripotency and, depending on their environment, can differentiate into dendritic cells or macrophages with specific cellular functions. They have high endocytic capacity and can be recognized by the expression of cell markers on the membrane, such as CD14+.

### 1.7. Stromal Cells

Stromal cells are a type of cell that make up certain supporting tissues surrounding other tissues and organs.

### 1.8. Histological Samples

Histological samples obtained from biopsies represent a confirmatory diagnostic method for various pathological conditions. Among these, samples obtained from solid tumors are indispensable for tumor classification and the choice of treatment. Among other variables observed in histological samples, it is possible to quantify the presence of cell populations associated with the immune response, such as lymphocytes, monocytes, and neutrophils. Additionally, tumor and stromal cells are often present alongside necrotic cells, which provides information about the activity of the immune response and its balance established with the tumor.

The sample preparation consisted of millimeter-thick sectioning of a previously paraffin-embedded piece of tissue, which was then placed on a glass slide. Subsequently, the sample was stained with hematoxylin-eosin, resulting in clear visualization of the morphological structures of the cells, such as the nucleus and cytoplasmic form and distribution (**Figure 1**). Subsequently, the different cell types are quantified either automatically or under the supervision of a pathology specialist.

**Figure 1:**
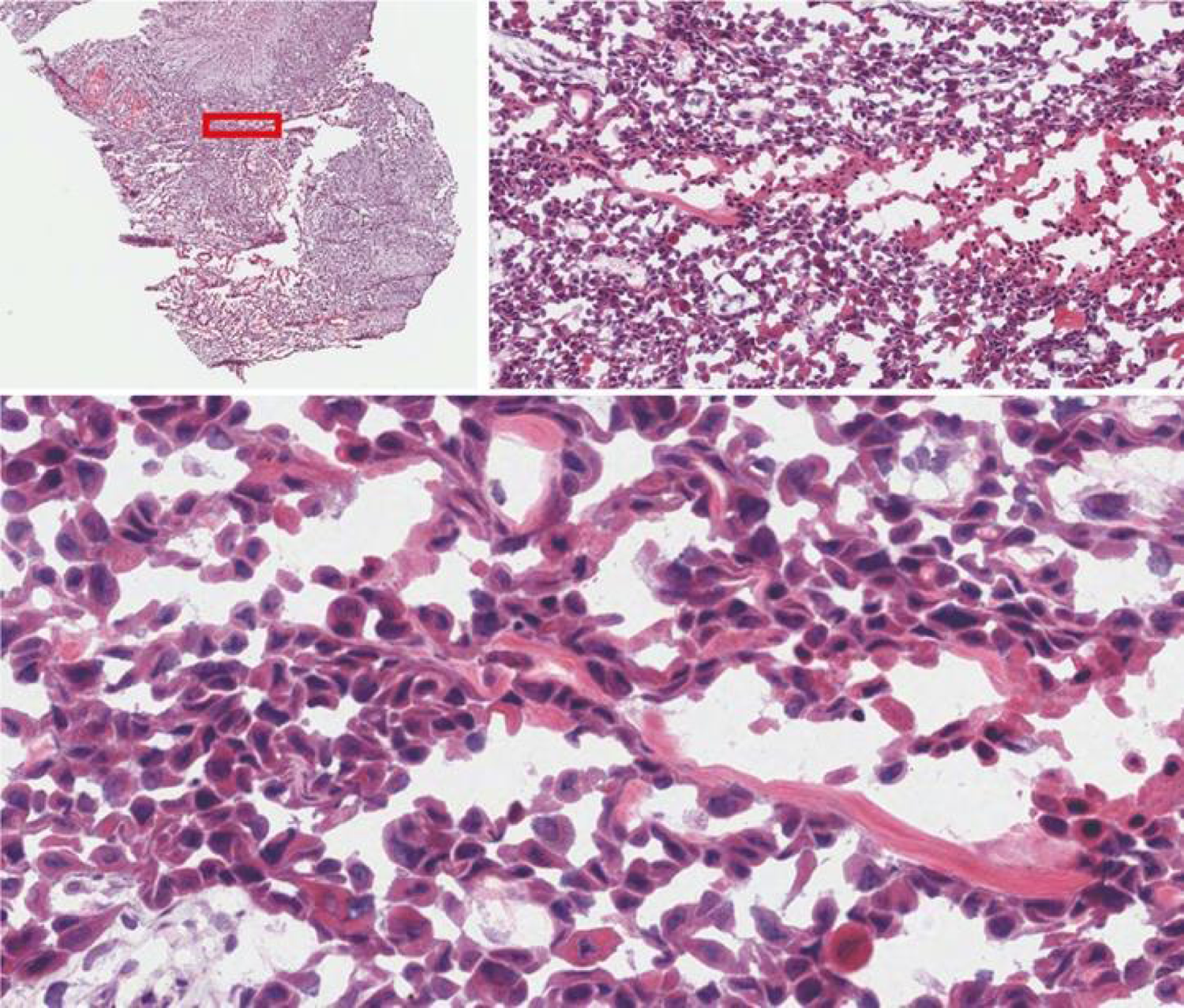
Histological sections with histochemical staining. From left to right and top to bottom, the complete analyzed tissue section is observed, followed by two consecutive magnifications performed in the region of the red rectangle. At maximum magnification, corresponding to the inset in the lower panel, cell nuclei are distinguished in dark blue, cellular cytoplasm in intense pink, and connective tissue as portions of light pink. The image shows a biopsy of a woman diagnosed with squamous cell carcinoma (slide ID:4b828b23-765f-4539-a55e-3f81de6e3907). It contains 15% stromal cells, 10% necrotic cells, and 75% tumor cells.

## 2. Results

### 2.1. The average survival in days of the analyzed patients is less than 4 years

Based on the analysis of the numerical variables, as shown in Figure 2, the average survival in days of the analyzed records was less than 1500 days, approximately four years (pink boxplot in the lower-left graph). Survival rates vary significantly based on the type of tumor and early diagnosis, and are also influenced by the existence and effectiveness of available treatments, among other genetic and environmental factors.

**Figure 2:**
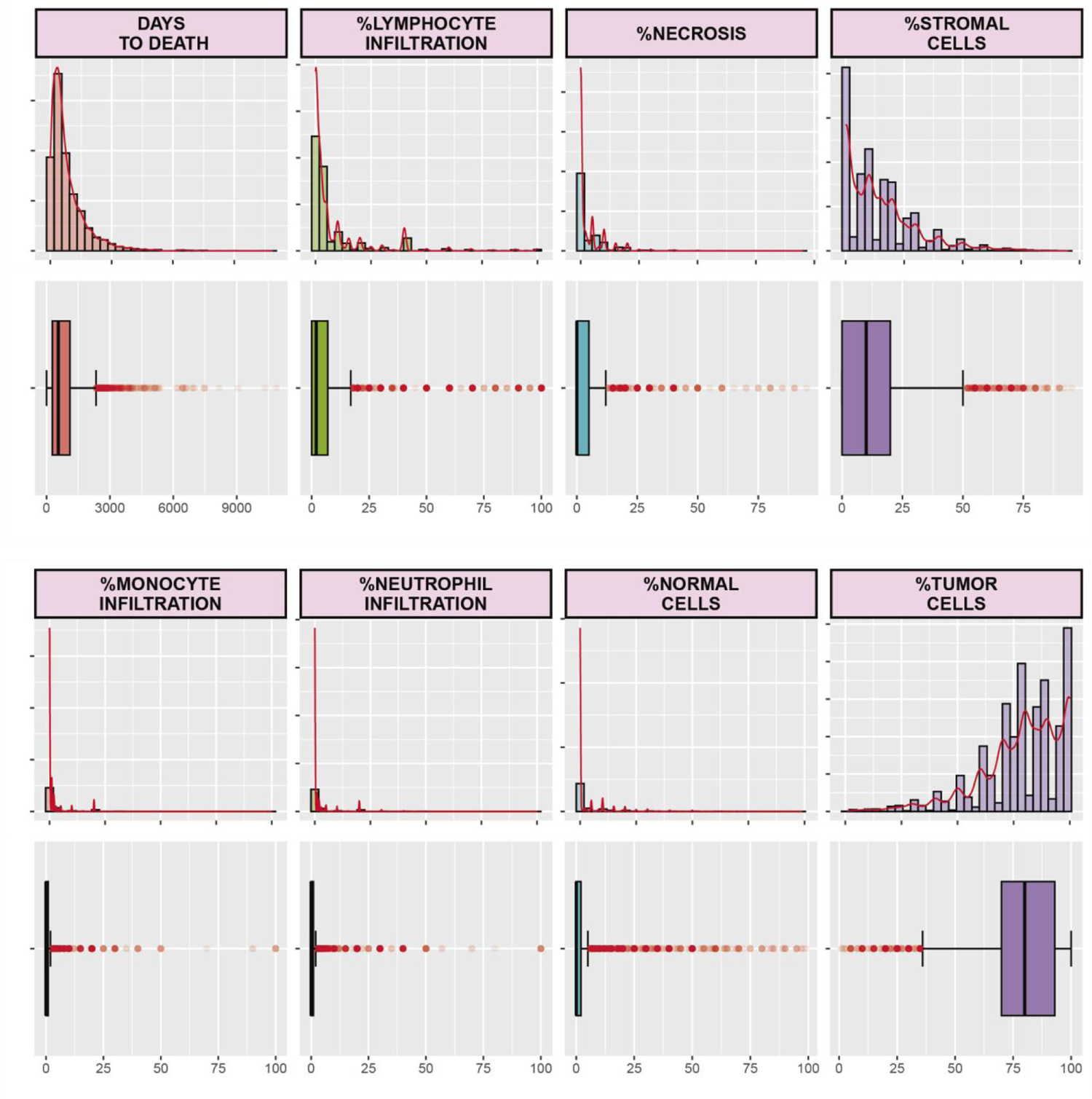
Distribution of Numerical Variables. Histogram and density plot accompanied by a box plot diagram for each numerical variable. From top to bottom and left to right, the values of the following variables are shown: survival in days following diagnosis (DAYS TO DEATH), percentages of infiltrating lymphocytes (%LYMPHOCYTES), percentage of necrotic cells (%NECROSIS), percentages of present STROMAL CELLS (%STROMAL CELLS), percentages of INFILTRATING MONOCYTES (%INFILTRATING MONOCYTES), percentage of INFILTRATING NEUTROPHILS (%INFILTRATING NEUTROPHILS), percentages of NORMAL CELLS (%NORMAL CELLS), and percentages of TUMOR CELLS (%TUMOR CELLS).

It is important to note that our survival analysis may be biased towards more severe cases of the disease, where the 5-year survival rates are low. This could be because the majority of missing data for this category correspond to individuals who have overcome the disease and are still alive.

Another interesting finding from the exploration of numerical variables is the specular distribution of all analyzed values and that of tumor cells. While the proportion of values related to tumor cells had an average above 75%, for all other quantified values, the mean distribution was below 25%. Regarding the analysis of the distribution of the numerical values, all corresponding histograms showed a unimodal structure, suggesting the existence of a single population for each variable.

Strikingly, in Figure 2, for tumor and stromal cells, the histograms display some reduced intermediate values (upper and lower right histograms in violet). This indicated the presence of related cell proportions that are infrequently observed.

### 2.2. The percentage of stromal and tumor cells is distributed in a contrasting manner

Similar to the rest of the analyzed numerical variables, the distribution of tumor cells was contrasting, showing higher average values than stromal cells. In particular, the shape of the histogram distribution for the percentage of stromal cells was an inverted reflection of the distribution of tumor cell percentage values (Figure 2, upper and lower right histogram graphs in violet).

As the tumor progresses, enzymes that destroy the structure of neighboring tissues are secreted. This hinders containment of the tumor mass and allows for physical expansion in the body. The ability of a tumor to degrade the supporting tissue is a prerequisite for tumor expansion (metastasis) to other regions of the body. The destruction of connective tissue implies the disappearance of stromal cells that constitute it. This was reflected in the specular distribution between the percentages of stromal and tumor cells.

### 2.3. The distribution of the primary diagnosis category shows differences in relation to the TP53 gene alteration

Similar to the exploration conducted for the numerical variables, we studied the distribution of the classes of categorical variables considered. For the variable “survival at one year,” most of the records did not have a value, so they were assigned the value “unknown” (Figure 3A). This category likely includes records of patients who are still alive (at the time of this analysis) after diagnosis.

**Figure 3:**
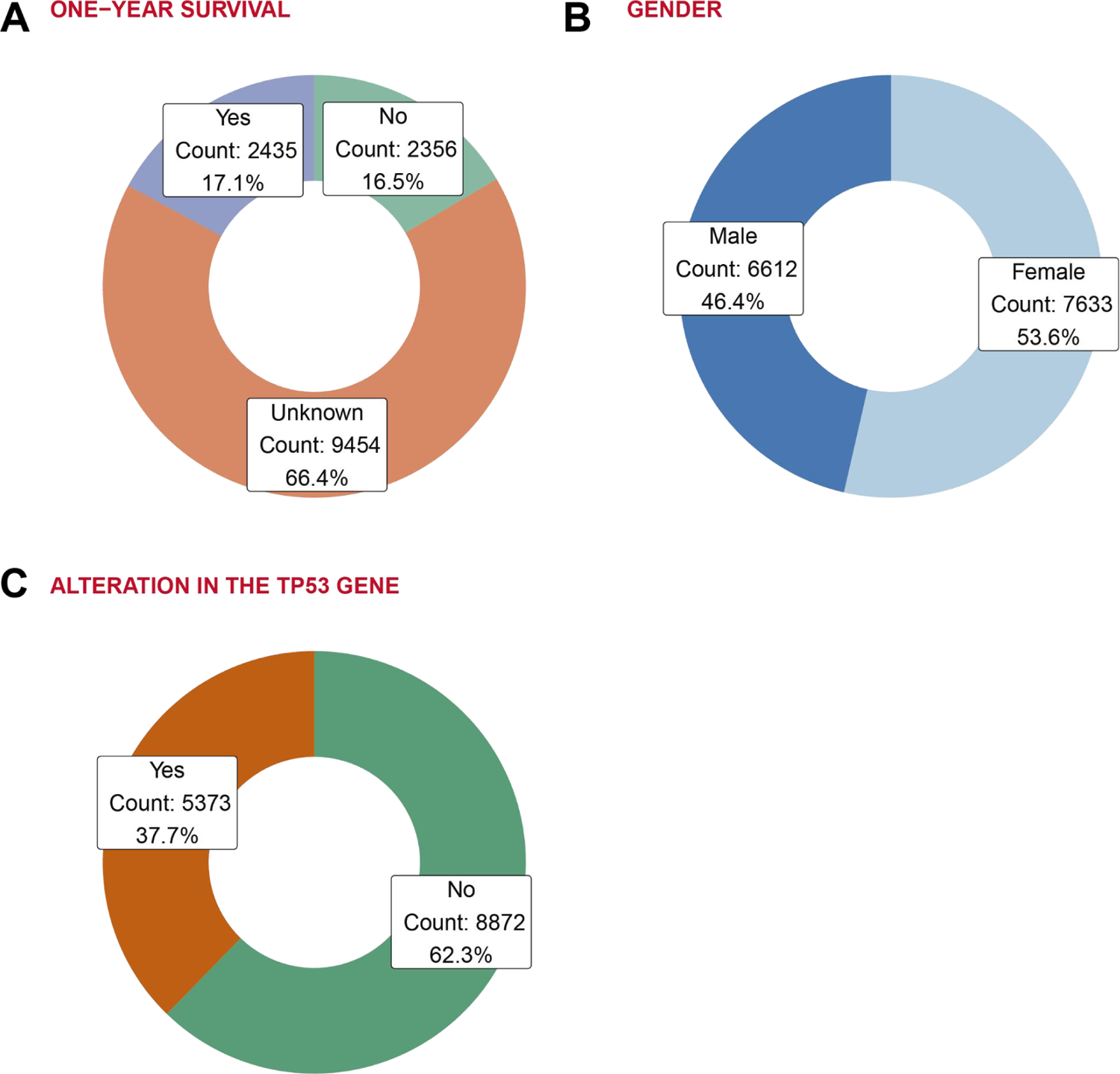
Total Count and Percentage of Categorical Variables Included in the Analysis. (A) Proportion and total count of records with survival exceeding one year after the primary diagnosis. (B) Proportion and total count of records for the binary variable sex. (C) Proportion and total number of patients with alterations in TP53 gene genotypes.

On the other hand, deaths occurred before one year accounted for 2356 records, while records where life exceeded one year totaled 2435 (Figure 3A). For the gender variable, the classes “male” and “female” are balanced, with 6612 and 7633 cases, respectively (Figure 3B).

Regarding the proportion of records with TP53 gene alterations, 62.3% did not have alterations, surpassing the class with alterations of 37.7% (Figure 3C). As TP53 alteration is the most frequent mutation found in tumors, one might expect values closer to 50% of the records. The discrepancy between the observed and expected data may be explained by the tumor types that are more likely to be included in the TCGA dataset.

As for the categories present in the primary diagnosis variable, 137 types of tumors were identified. The number of categories per case was concentrated in seven tumor types (62.6% of the total records), whereas the rest of the tumor types did not exceed 5% of the total records (Figure 4A). The most common tumor types were adenocarcinoma (16.2%), Squamous Cell Carcinoma (10.6%), Invasive Ductal Carcinoma (9.4%), Serous Cystadenocarcinoma (8%), glioblastoma (7.4%), Clear Cell Adenocarcinoma (5.7%), and Papillary Adenocarcinoma (5.3%).

**Figure 4:**
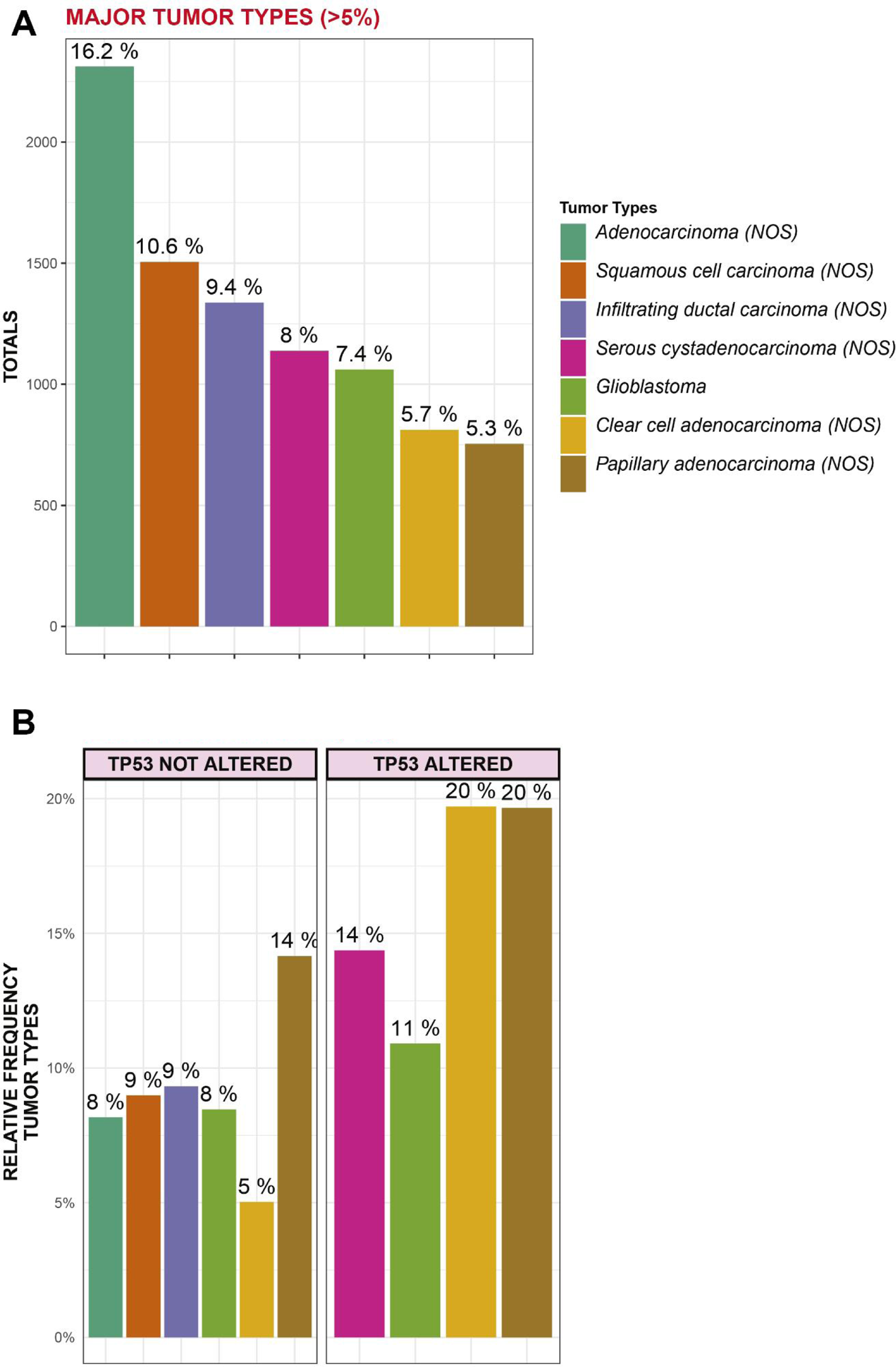
Major Tumor Types. (A) The proportion of patients according to the primary diagnosis type for cases that exceeded 5% of the total. (B) Relative frequency of patients with or without alterations in TP53 gene according to the primary diagnosis type for cases that exceeded 5% of the total.

The distribution of tumor types among patients with mutated TP53 and those with unaltered TP53 was explored. Contrary to this hypothesis, the majority distribution of tumor types (> 5%) present in both sets was different (Figure 4B). For example, while Serous Cystadenocarcinoma (NOS) represented 14% of the total mutated cases, it did not exceed 5% of the unaltered cases. Conversely, Invasive Ductal Carcinoma (NOS) did not exceed 5% of the mutated cases but represented almost double (9%) of the cases without alterations in the TP53 gene. Another extreme example of this imbalance in the number of mutated and unmutated tumors is Clear Cell Adenocarcinoma (yellow bar, 20% and 5%, respectively). These asymmetries may reflect unknown underlying molecular mechanisms.

### 2.4. The presence of neutrophils, monocytes, and lymphocytes correlates positively with each other

Continuing with the analysis of the relationships between variables, principal component analysis was performed based on immunological numerical values. Figure 5A shows the biplot graph of the decomposition into two principal components that together explain 57.3% of the observed variability (PC1=31.9 and PC2=25.4). In this graph, the red dots correspond to all the records projected into a lower-dimensional space (dim=2). The black arrows symbolize the contribution and direction of the variables corresponding to the projection axes. The values of monocytes, neutrophils, and lymphocytes contributed similarly and in the same direction (perpendicular to the other variables). On the other hand, coinciding with what was observed in Figure 2, tumor cells influence stromal cells and normal cells in the opposite direction.

**Figure 5:**
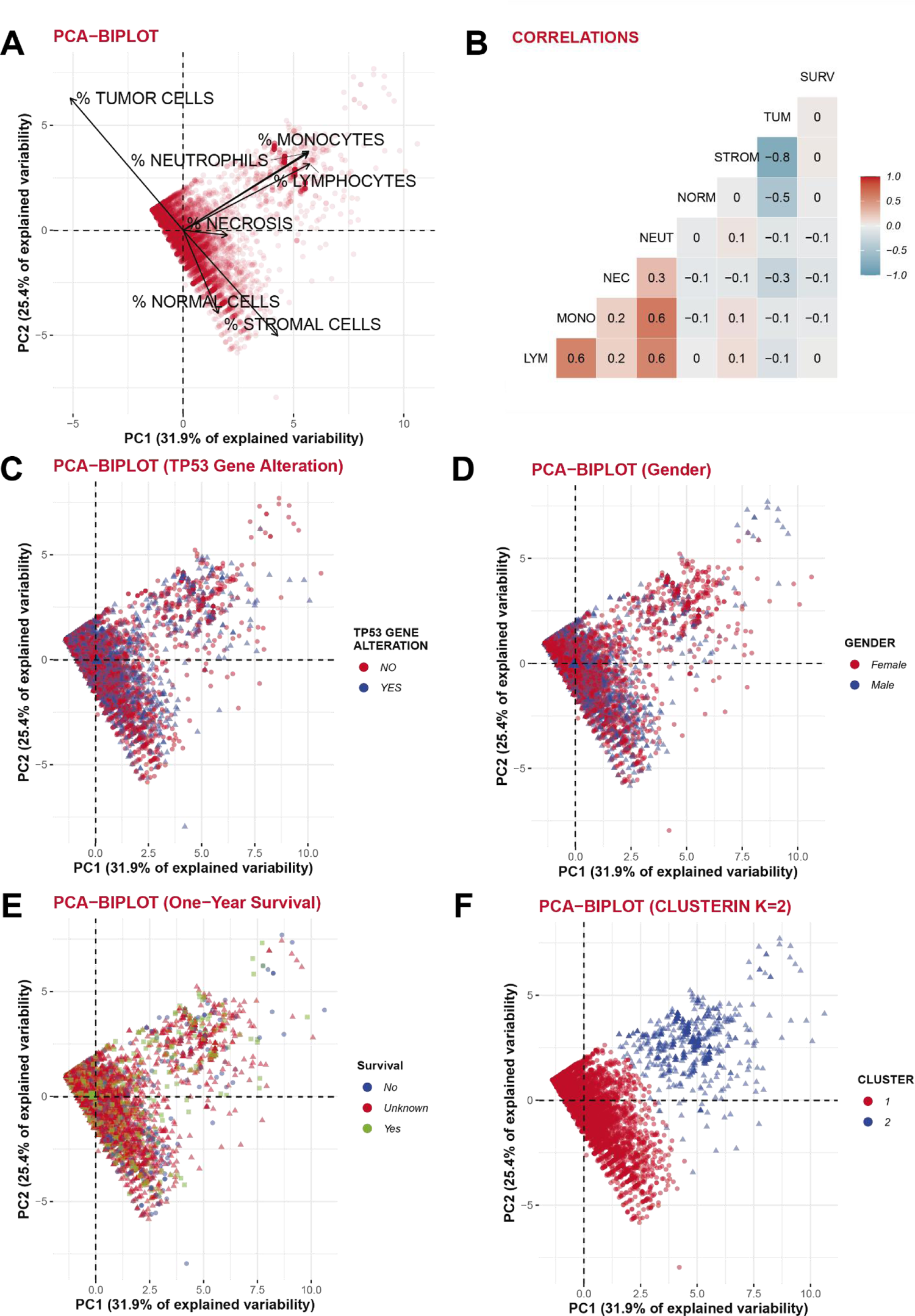
Correlation and clustering of numerical variables. Scatter plots corresponding to dimensionality reduction using Principal Component Analysis (PCA) are shown for the first two components (A, C, D, E, F). (A) PCA scatter plot displaying the direction and contribution of the variables to the projection on the axes. (B) Correlation plot of numerical variables. Strong negative correlations are shown in deep blue, whereas strong positive correlations are indicated in red. (C) PCA plot where individuals with (red) and without (blue) TP53 gene alterations are visualized. (D) PCA plot distinguishing female individuals (red) and male sex (blue). (E) PCA plot showing individuals whose survival surpasses (green) or does not surpass (blue) a certain threshold, with unknown survival shown in red. (F) PCA plot representing the cluster membership of records generated by unsupervised clustering (k-means). Cluster 1 is represented in red, and cluster 2 is shown in blue.

The correlation graph between the variables (Figure 5B) quantifies the positive and negative associations between variables. For example, the percentage of stromal cells was negatively correlated with the percentage of tumor cells (cor=-0.8). Contrary to this hypothesis, the presence of necrotic tissue in the sample did not correlate with survival (Days to death), but it did correlate slightly with the percentage of neutrophils (cor=0.3). Detection of neutrophils in necrotic tissues is a favorable prognostic indicator for some types of tumors ^8^.

Figure 5A also shows the presence of two groups of records. One group had a high percentage of infiltrating monocytes, neutrophils, and lymphocytes, whereas the other group had a reduced percentage of these cells. However, these groupings did not seem to concentrate a differential distribution of records in terms of sex (Figure 5D), one-year survival (Figure 5E), or the presence of TP53 gene alterations (Figure 5C). This visual clustering was also consistent with unsupervised clustering (k-means clustering, k=2) in Figure 5F, where the optimal number of clusters was determined by the maximum silhouette.

### 2.5. The unsupervised clustering using the K-means and DBSCAN algorithm generates two groups that differ in the proportion of immune system cells

The distribution of each numerical variable was considered for each group based on the clusters determined by unsupervised clustering using the K-means algorithm. Figure 6A shows boxplot graphs representing the distribution of each numerical variable for each cluster (cluster 1 in green and cluster 2 in orange). The distribution between the two clusters showed differences only for the variables “lymphocyte infiltration”, “monocyte infiltration” and “neutrophil infiltration”.

**Figure 6:**
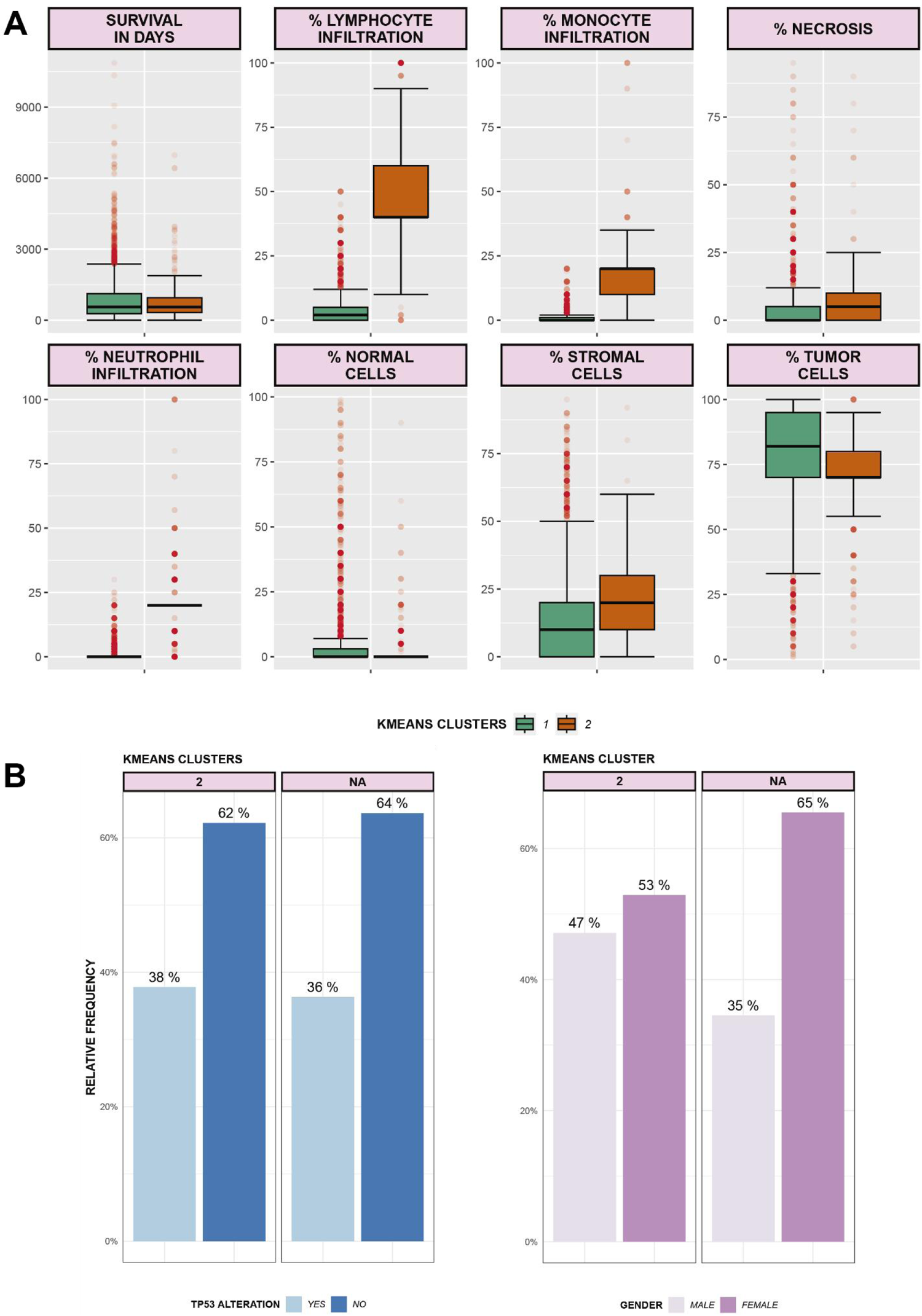
Cluster composition determined by unsupervised clustering. (A) Boxplot of each numerical variable for both clusters. (B) Proportion of the number of classes for the variables TP53 gene alteration (blue) and gender (pink) among the clusters determined by k-means (k=2).

In addition, according to the analysis conducted, the proportion of classes in the variables “gender” and “alteration in the TP53 gene” did not show differences between the clusters (Figure 6B).

### 2.6. DBSCAN clusters didn’t show differences in terms of the proportion of classes for the variable “one-year survival”

Because the visualization of the data suggested the existence of two clusters with different densities (coinciding with the clustering generated by the K-means algorithm), DBSCAN clustering was employed (Figure 7A). For these new clusters, no differences were found in terms of the proportion of classes for the variable “one-year survival” (Figure 7B). However, DBSCAN revealed again that the two groups differed in the proportion of immune cells present in the samples. This indicates that the composition of immune cells may play a role in the clustering of samples and could be related to the underlying biology and characteristics of the tumors.

**Figure 7:**
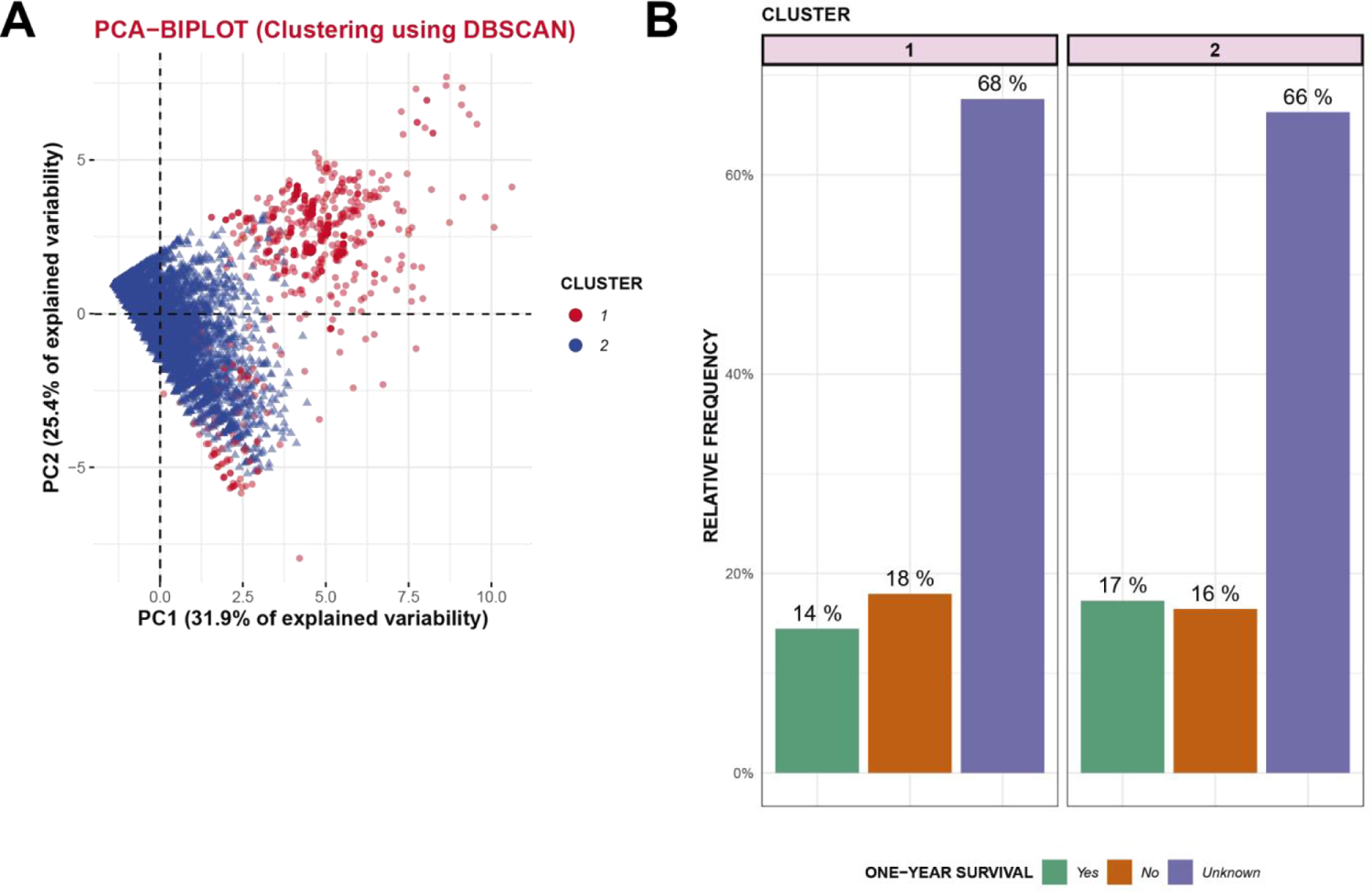
DBSCAN Clusters. (A) PCA plot showing the cluster membership of the records generated by unsupervised clustering using DBSCAN. Cluster number 1 is represented in red, and cluster number 2 is shown in blue. (B) Proportions of classes for the variable “survival at one year.” The colors represent different categories: green for “YES,” brown for “NO,” and violet for “unknown.” These proportions are shown for the clusters determined using DBSCAN.

Similar to the analysis conducted for the clusters generated by K-means, the distribution of numerical variables for the new DBSCAN clusters was studied (Figure 8A). Differences were found in the distribution of the variables “percentage of infiltrating lymphocytes,” “monocytes,” and “neutrophils.” This result is consistent with that observed in Figure 5A, where these variables contributed to the direction of Cluster 2 in Figure 8A.

**Figure 8:**
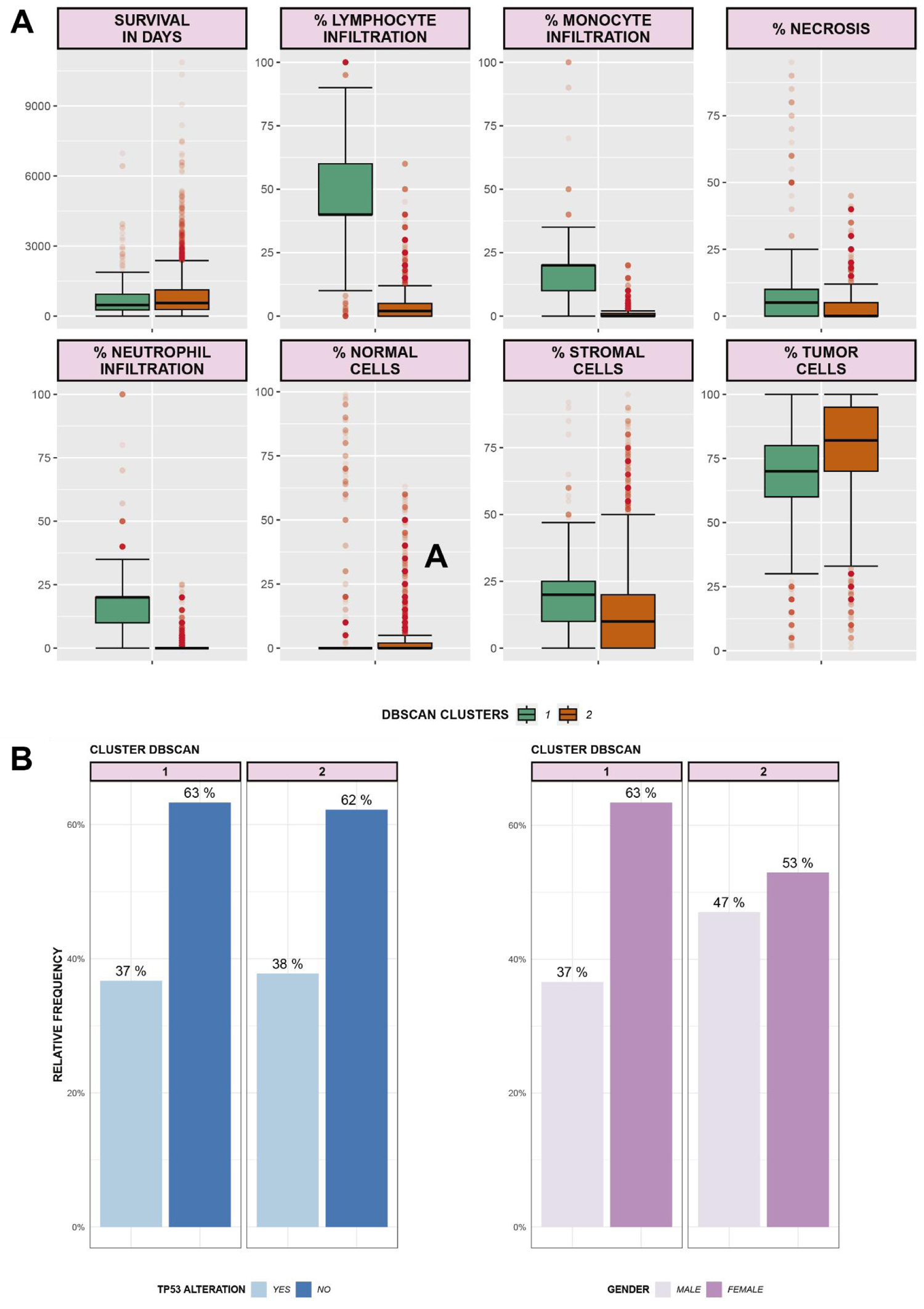
Clusters determined by unsupervised DBSCAN clustering. (A) Boxplot of each numerical variable for both clusters. (B) Proportion of the number of classes for the variables “TP53 gene alteration” (blue colors) and “gender” (pink) among the clusters determined by DBSCAN.

Regarding the analysis of proportions for the categorical variables “gender” and “presence of alterations in TP53,” cluster 2 showed an alteration in the gender frequency compared to cluster 1 (Figure 8B). Cluster 1 had a higher number of female records (37:63, male:female), while cluster 2 had more balanced proportions (47:53, male:female). These findings suggest that the composition of immune cells and sex may be important factors for distinguishing the two clusters generated by DBSCAN.

### 2.7. Classification models

Analysis of the supervised classification model using logistic regression revealed that certain tumor types, specifically Malignant Melanoma, Ovarian Cancer, and Glioblastoma, have a significant impact on patient survival. These tumor types were identified as the most important predictors of the model, indicating that they were associated with poorer survival outcomes. This finding is consistent with the aggressive nature and lower survival rates of these types of tumors.

Among immunological variables, the percentage of necrosis was the most important predictor of patient survival. This result supports the initial hypothesis that a higher percentage of necrotic tissue negatively affects survival. However, it is worth noting that the correlation between the percentage of necrosis and survival was not significant (Figure 5B).

### 2.7. Lasso Model

Based on the tumor classes selected by the Lasso method, a new categorical variable was constructed, grouping all tumor types into four classes: Malignant melanoma, Serous Ovarian Cystadenocarcinoma, Glioblastoma, or Other. Additionally, the data were divided into training (training) and testing (test) sets, where the predictive class “Survival at one year” was balanced (**Supplementary** Figure 2).

The proposed models for logistic regression were as follows:

However, the worst model coincided with the single normal cell model. As observed in the corresponding ROC curves for both models (**Figure 10A**), for the worst model, the prediction probability remained close to 50% throughout, which is equivalent to a random chance (binary variable 50%-50%).

**Figure 9:**
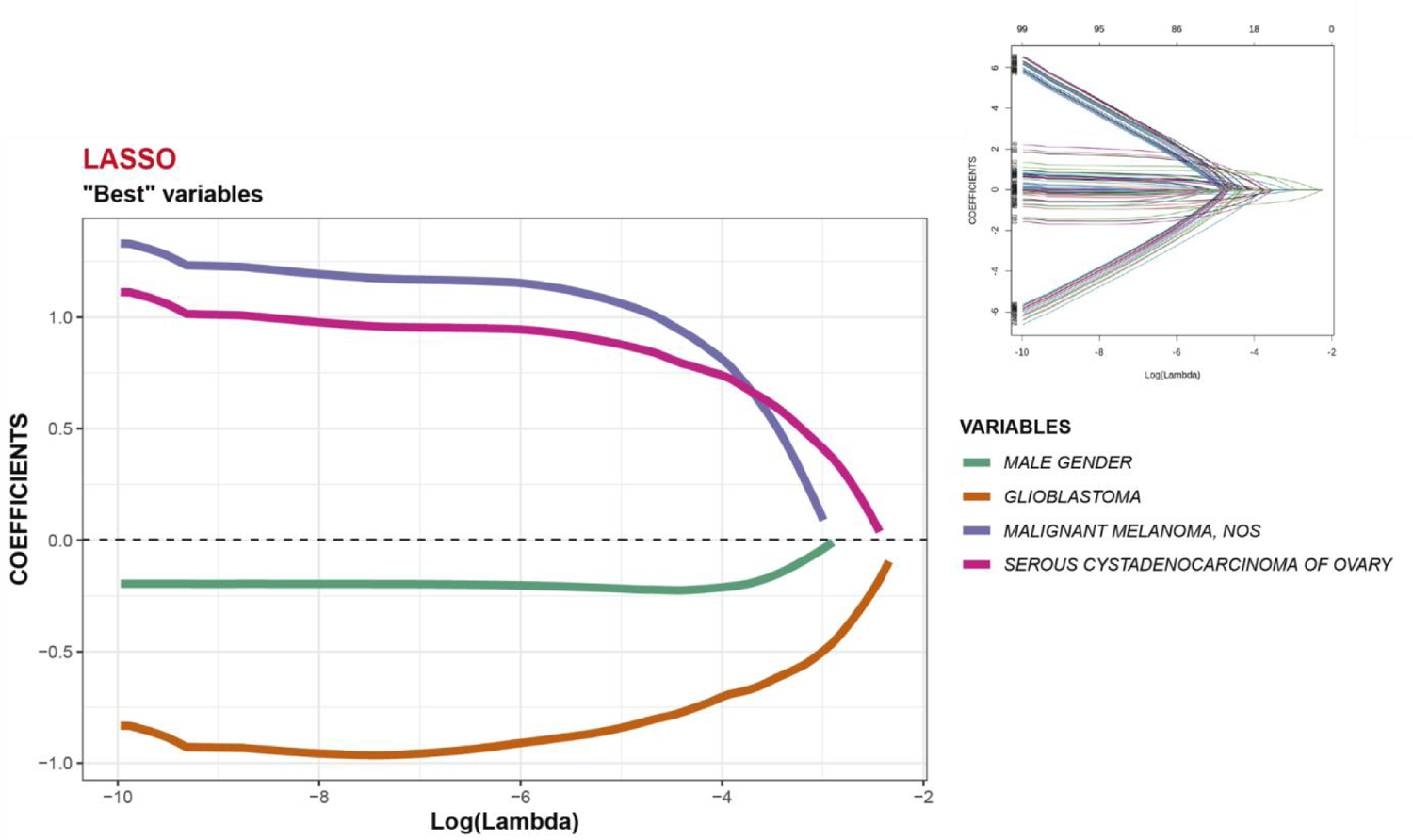
Generalized Linear Model. Variable selection using the LASSO method. In the upper right corner, all considered variables are shown, as well as how they survive under incremental lambda penalties. On the right side, the magnification is presented based on the upper graph.

**Figure 10:**
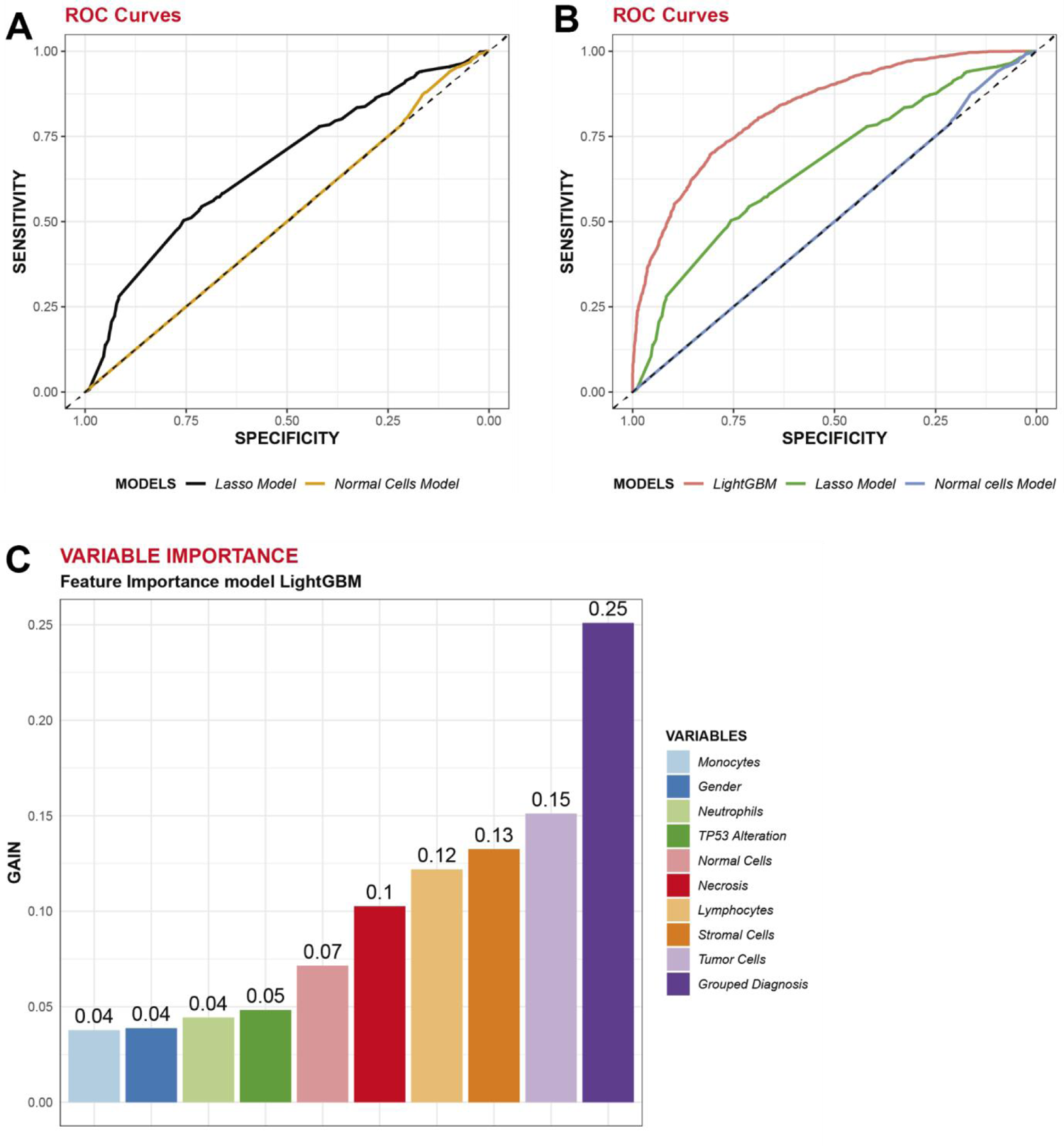
Metrics. (A) ROC curves for the LASSO model and the normal cell model. (B) ROC curves for the Lasso, normal cell, and LightGBM models. (C) Bar graph showing variable gains according to the LightGBM model.

Finally, according to this approach, the primary diagnosis type among the grouped diagnostic categories is of greater relevance in terms of one-year survival probability. Consequently, the impact of immune variables on prediction was lower.

### 2.8. LightGBM Classifier

In the last part of this study, a classification model based on decision trees (LightGBM model) was trained to predict one-year survival. The purpose of this training is to determine the relative contributions of the attributes considered by the algorithm. Variable gains are obtained when they are used as splitters in the decision tree. In both the linear regression and LightGBM models, the data were trained on the same training set and applied in both cases to the same testing set. As observed in Figure 10C, similar to what was detected by the Lasso model for variable selection, the most relevant attribute for the tree was Grouped Diagnosis (gain of 0.25). In contrast to the LASSO model, there are many immune variables with similar gains that are relevant for the classifier (tumor cells 0.15, stromal cells 0.13, lymphocytes 0.12, and necrotic cells 0.1).

Figure 10B shows the overlap of the ROC curves for the Lasso model, single normal cell model, and LightGBM model. The classifier model has a larger area under the curve than the lasso model, even without parameter optimization (default LightGBM). This result can be interpreted as an advantage of the decision tree model derived from the utilization of all attributes present in the training and testing sets. In contrast, the Lasso model only considers the variables proposed for regression (in this case, grouped diagnosis, sex, and percentage of necrotic cells). It is important to mention that in **Figures 10A** and **10B**, the ROC curves were constructed based on the application of the models to the testing set.

## Materials and methods

### 3.1. Data

#### 3.1.1. Clinical, pathological, and genetic information

The database was constructed from clinical information and pathological information from 10,528 patients with various types of cancer available in The Cancer Genome Atlas (TCGA) - Cancer Genome Atlas database. The following numerical variables were selected: survival in days, percentage of infiltrating neutrophils, percentage of infiltrating lymphocytes, percentage of tumor cells, percentage of stromal cells, percentage of normal cells, and percentage of infiltrating monocytes in different pathological samples. Additionally, the categorical variables of sex, presence or absence of alterations in the TP53 gene, and primary diagnosis were selected.

#### 3.1.2. Specific mutations

Specific information on the presence or absence of mutations in the coding sequence of the p53 protein (TP53 gene) was incorporated. The corresponding information was generated using cBioPortal (https://www.cbioportal.org/), selecting cases belonging to TCGA studies, and subsequently performing a merge using the patient code as the intersection attribute between the tables.

### 3.2. Methodology

Both preprocessing and subsequent analyses were performed using R-Studio. In summary, the numbers of lymphocytes, neutrophils, monocytes, normal cells, stromal cells, and tumor cells were obtained from the pathological samples (slides) associated with the patients. The same tissue also provided DNA and RNA for further analysis after review by histopathology specialists. Given the existence of more than one histological sample per patient, only those samples that presented with tumor cells were considered, resulting in the following number of slides per patient (**Table 2**).

**Table 1:** Models based on the Deviance calculated on the training dataset.

| Ord. | Deviance | Models |
| --- | --- | --- |
| 1 | 4.348,51 | Grouped Diagnosis + Gender + Percentage of Necrosis |
| 2 | 4.370,53 | Grouped Diagnosis |
| 3 | 4.575,77 | Gender |
| 4 | 4.597,89 | %Neutrophils + %Lymphocytes + %Necrosis + %Monocytes |
| 5 | 4.601,39 | %Neutrophils + %Lymphocytes + %Necrosis |
| 6 | 4.601,64 | Necrosis |
| 7 | 4.641,35 | %Tumor Cells + %Normal Cells |
| 8 | 4.642,32 | Tumor Cells |
| 9 | 4.644,06 | Monocytes |
| 10 | 4.645,48 | Stromal Cells |
| 11 | 4.647,09 | Neutrophils |
| 12 | 4.647,22 | Lymphocytes |
| 13 | 4.647,34 | TP53 Alteration |
| 14 | 4.647,41 | Normal Cells |

**Table 2:** Total and percentage distribution of the number of samples per patient.

| Number of samples per patient | Number of patients | Percentage |
| --- | --- | --- |
| 1 | 7099 | 67,43 |
| 2 | 3253 | 30,9 |
| 3 | 98 | 0,93 |
| 4 | 59 | 0,56 |
| 5 | 12 | 0,11 |
| 6 | 4 | 0,04 |
| 8 | 2 | 0,02 |
| 10 | 1 | 0,01 |
| **TOTAL** | 10528 | 100 |

Based on this selection, each unique triplet of patients and samples was considered as an individual record to increase the amount of available data (data augmentation) for subsequent analysis. The final number of records considered amounted to 14,245 (an increase of 35.3%). The scripts used for data pre-processing and construction of each figure are available in the following repository:

### 3.3. Missing Attributes

The categorical variables, sex, primary diagnosis, and TP53 gene alteration, did not have any missing attributes. The distribution of missing values for the associated numerical variables is presented in **Table 3**.

**Table 3:** Total and percentage distributions of the number of missing attributes for each variable in the total records.

| Variables | Missing values | Percentage |
| --- | --- | --- |
| Survival (in days) | 9454 | 66,4 |
| TP53 alteration | 0 | 0 |
| Percentage of tumor cells | 0 | 0 |
| Percentage of stromal cells | 79 | 0,6 |
| Percentage of infiltrating neutrophils | 5397 | 37,9 |
| Percentage of normal cells | 111 | 0,8 |
| Percentage of infiltrating monocytes | 5418 | 38 |
| Percentage of necrotic cells | 4 | 0 |
| Percentage of infiltrating lymphocytes | 4992 | 35 |
| Primary diagnosis | 0 | 0 |
| Gender | 0 | 0 |

For the construction of predictive and classification models, only complete records for the variable “survival in days” were considered (a total of 4791 records). The missing values for immunological variables (percentage of stromal cells, infiltrating neutrophils, normal cells, infiltrating monocytes, and infiltrating lymphocytes) were imputed with zeros. Duplicate data points were eliminated from the dataset.

### 3.4. Variable Generation

A categorical variable named survival was created based on the data from “Survival” was created based on the data from “survival in days” (numerical). It consisted of “YES” (2435 records) if the patient’s survival was longer than one year after the primary diagnosis year, or “NO” (2356 records) if it was not longer. For records where “survival in days” had missing values, the “Survival” variable was labeled as “Unknown” (9454 records).

### 3.5. Supervised Classification

The categorical variable “Survival” was chosen as the predictor variable. Only cases with a value for the numerical variable “survival in days” were considered (n = 4791). For these cases, missing numerical values were imputed to zero, which is biologically relevant to histological samples. As mentioned before, the sex and primary diagnosis categories had no missing values.

### 3.6. Normality and Model Selection

The numerical variables did not follow a univariate normal distribution and the assumption of multivariate normality was rejected. Therefore, only logistic regression models were used, with the predictor variable being “Survival at one year” (where the category “YES” was coded as “1” and the category “NO” as “0”).

### 3.7. Unsupervised Classification

For unsupervised clustering, scaled numerical variables (percentage of infiltrating monocytes, lymphocytes, neutrophils, normal cells, stromal cells, tumor cells, and necrotic cells) were used. Similar to the models mentioned above, the missing values for these variables were imputed to zero. Two clustering algorithms were employed.

First, the k-means algorithm is used as an unsupervised clustering method. The optimal number of clusters for this algorithm was determined by studying the silhouette scores for different values of k in the range [1, 15]. The analysis showed that optimal separation was achieved for k = 2.

Second, the DBSCAN algorithm was used with the following parameters: eps = 2.7, Min Pts = 700.

### 3.8. lightGBM Classifier

For comparison with the regression model, the “Grouped Diagnosis” variable generated from the Lasso model was used. The variables “survival in days,” “id,” and “primary diagnosis” primary diagnoses were excluded from the training and test sets. A lightGBM model without parameter optimization (default: objective= “binary”) was used.

## 4. Conclusions

Based on the comprehensive analysis conducted in this study, we have derived several significant conclusions that shed light on the factors influencing the one-year survival probability of patients with cancer.

Our findings indicate that the mean survival time for the analyzed patients was < four years. This suggests that overall survival in this cohort is limited and underscores the need for improved prognostic models and therapeutic interventions.

Contrary to our hypothesis, the presence of immune cells did not have a significant impact on patient survival. This intriguing result may warrant further investigation into the complex interplay between the immune system and tumor progression in this specific context.

The percentage of necrotic tissue was identified as an essential variable in generating a predictive logistic regression model and featured as a significant attribute for the decision tree. Although it did not directly correlate with patient survival time, it proved to be a critical factor in predicting survival outcomes, suggesting its relevance in tumor behavior.

Our analysis revealed heterogeneous alterations in the TP53 gene among different tumor types. This diversity in TP53 mutations suggests that the role of this gene in tumorigenesis and prognosis may vary depending on tumor context.

Specific primary diagnoses, such as Malignant Melanoma, Serous Ovarian Cystadenocarcinoma, and Glioblastoma, have emerged as strong determinants of the one-year survival probability. Patients diagnosed with these aggressive tumor types experienced a significantly reduced probability of survival beyond one year, emphasizing the clinical importance of early diagnosis and targeted treatment strategies.

Finally, the decision tree classifier emphasized the importance of certain immune variables, including tumor cells, stromal cells, lymphocytes, and necrotic cells, in predicting survival outcomes. This underscores the relevance of the tumor microenvironment and immune response in influencing patient survival.

In conclusion, our comprehensive investigation provides valuable insights into the multifaceted factors impacting one-year survival of patients with cancer. These findings contribute to our understanding of tumor biology, immune responses, and clinical outcomes. Further research on these results could lead to the development of more accurate prognostic models and targeted therapies, ultimately improving patient outcomes for specific tumor types.

## 5. Discussion

The proposed models provided the main determinants of the probability of one-year survival in diagnosed individuals. Particularly, the most relevant observations regarding tumor types align with some of the most aggressive neoplasms and partially confirm, as in the case of glioblastoma, findings from similar studies ^9^.

Additionally, a conditional distribution of tumor classes with or without alterations in TP53 was observed. For example, ovarian cancer has the highest mutation rate^10,11^, which is consistent with some of the results shown in Figure 4B. Another relevant observation generated from this study was the detection of two clusters among the records that differed in the presence of lymphocytes, monocytes, and neutrophils. Although these clusters did not show differences in terms of survival, other studies have correlated infiltration levels negatively or positively with overall survival, depending on the tumor type^12^.

Furthermore, regarding immune variables, both the Lasso model and LightGBM classifier emphasized the importance of the percentage of necrosis. Generally, the presence of necrotic cells in tumor regions reflects the existence of hypoxia, which may indicate more aggressive tumor behavior ^13,14^.

Finally, the results obtained find parallels and support many other observations made in the field and present novel aspects for future initiatives and working hypotheses.

## Data Availability

All data produced in the present study are available upon reasonable request to the authors

## Notes

### Competing Interest Statement

The authors have declared no competing interest.

### Funding Statement

This study did not receive any funding

### Author Declarations

The results shown here are in whole or part based upon data generated by the TCGA Research Network: https://www.cancer.gov/tcga.

