## Supplemental figures for "Immunological variables and tumor types influence one-year survival probability in cancer patients: A comprehensive analysis using logistic regression and decision tree models"

**SUPLEMENTARY DATA**

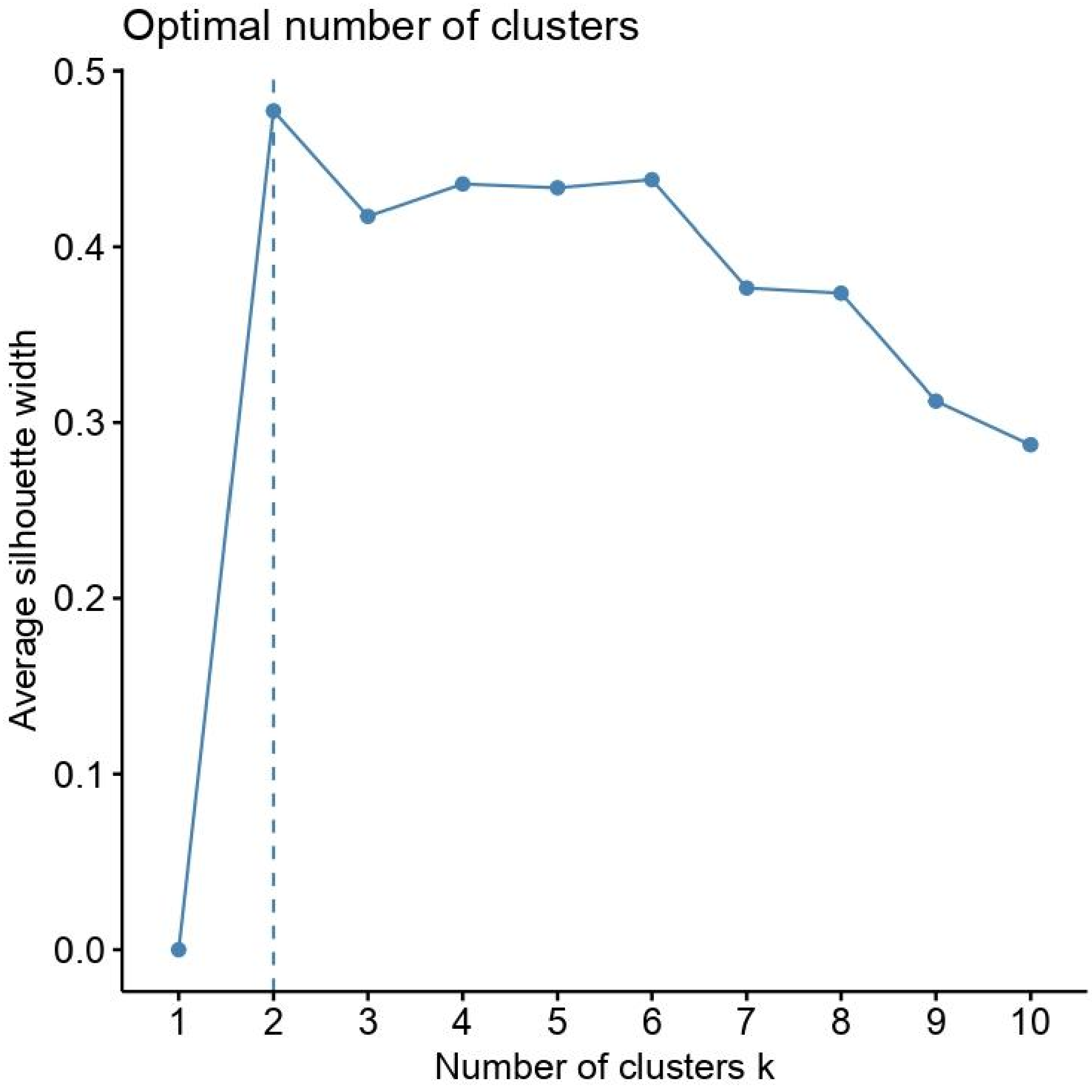


**Supplementary Figure 1:** Silhouette profile plot for different clustering values of k. The optimal value of K=2 is highlighted with a dashed dotted line.

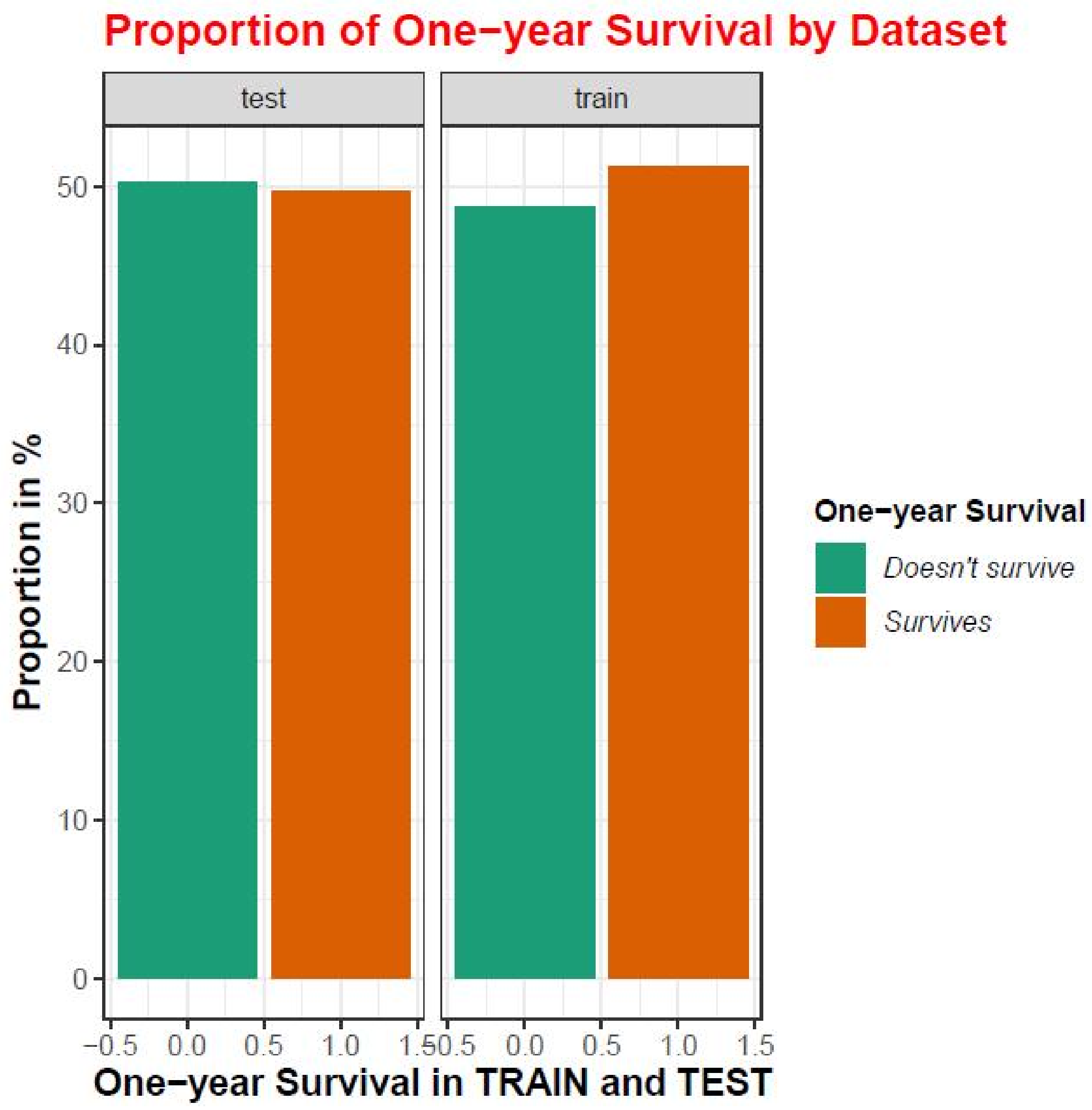


**Supplementary Figure 2:** Percentage distribution of the target variable 'survival' in the train and test datasets.

| Order | Term | Step | Lambda | Dev.ratio |
|---|---|---|---|---|
| 1 | primary_diagnosisGlioblastoma | 2 | 0,0961382595813188 | 0,0054580586870013 |
| 2 | primary_diagnosisSerous cystadenocarcinoma, NOS | 3 | 0,0875976015682608 | 0,0113295307816977 |
| 3 | gendermale | 8 | 0,0550138467454355 | 0,0345689758955007 |
| 4 | primary_diagnosisMalignant melanoma, NOS | 9 | 0,0501265682250862 | 0,0391562485162739 |
| 5 | primary_diagnosisClear cell adenocarcinoma, NOS | 11 | 0,0416159575706325 | 0,0467592116320845 |
| 6 | primary_diagnosisTransitional cell carcinoma | 14 | 0,0314809457432529 | 0,0565820442084601 |
| 7 | primary_diagnosisCarcinoma, diffuse type | 15 | 0,0286842652885445 | 0,0593568363798943 |
| 8 | primary_diagnosisLobular carcinoma, NOS | 16 | 0,0261360342174581 | 0,0620515027808881 |
| 9 | percent_necrosis | 17 | 0,0238141809715079 | 0,0648960074535377 |
| 10 | primary_diagnosisMesothelioma, biphasic, malignant | 17 | 0,0238141809715079 | 0,0648960074535377 |
| 11 | primary_diagnosisMucinous adenocarcinoma | 17 | 0,0238141809715079 | 0,0648960074535377 |
| 12 | primary_diagnosisAdenocarcinoma, intestinal type | 18 | 0,0216985947686321 | 0,0682597566471903 |
| 13 | primary_diagnosisInfiltrating duct carcinoma, NOS | 18 | 0,0216985947686321 | 0,0682597566471903 |
| 14 | primary_diagnosisOligodendroglioma, NOS | 18 | 0,0216985947686321 | 0,0682597566471903 |
| 15 | primary_diagnosisRenal cell carcinoma, chromophobe type | 18 | 0,0216985947686321 | 0,0682597566471903 |
| 16 | primary_diagnosisSignet ring cell carcinoma | 18 | 0,0216985947686321 | 0,0682597566471903 |

**Supplementary table 1:** Raw values of Lasso coefficients, sorted by descending relevance. The first numerical variable, percent_necrosis, associated with the histological sample, is highlighted in yellow.
